# Assessing the cost-effectiveness of economic strengthening and parenting support for preventing violence against adolescents in South Africa: an economic modelling study using non-randomised data

**DOI:** 10.1101/2023.02.09.23285712

**Authors:** William E. Rudgard, Sopuruchukwu Obiesie, Chris Desmond, Marisa Casale, Lucie Cluver

## Abstract

There is limited evidence around the cost-effectiveness of interventions to prevent violence against children in low- and middle-income countries. We developed a decision-analytic model to estimate the cost-effectiveness of 1) a community outreach intervention to link eligible households to anti-poverty cash grants, and 2) a group-based parenting support intervention, and 3) a group-based parenting support ‘plus’ linkage of eligible households to anti-poverty cash grants intervention for preventing adolescent violence victimisation in Mpumalanga province, South Africa.

The target population was families with an adolescent living below the national food poverty line. Modelled violence outcomes were emotional, physical, and sexual abuse. Intervention effectiveness was conditional on interventions’ effect on two evidence-based protective factors for adolescent violence: food security and caregiver supervision. Cost-effectiveness ratios were expressed per DALY averted and evaluated against a South Africa-specific willingness-to-pay threshold. We varied model parameters to consider routine service delivery versus trial-based costing, and population-average versus high prevalence of violence.

For routine service delivery costing, both community grant outreach and parenting support interventions were cost-effective at population-average prevalence (ACER=USD2,650, and ACER=USD2,830 per DALYs averted, respectively), and high prevalence of violence (ACER=USD1,330 and ACER=USD2,305 per DALYs averted, respectively). The incremental cost-effectiveness of adding grant linkage to parenting support was USD271 and USD177 at population-average and high prevalence of violence, respectively. For trial-based costing, none of the interventions were cost-effective at population-average prevalence of violence, and only community grant outreach was cost-effective at high prevalence of violence (ACER=USD2400 per DALY averted). Cost-effectiveness estimates are expected to be conservative based on our only modelling intervention effects on three violence outcomes via two protective factors.

Findings indicate that investments in community grant outreach, and parenting support interventions are likely to be cost-effective for preventing adolescent violence. Adding a grant linkage component to parenting support would enhance this approaches cost-effectiveness.

## Introduction

Annually, 1.4 billion children are estimated to be affected by violence worldwide, incurring more than 5.1 million disability-adjusted life years (DALYs) (1,2). To support countries in their efforts to reduce child violence the World Health Organization (WHO) and eight other global agencies have endorsed seven evidence-based strategies to address violence against children, known collectively as INSPIRE (3). These seven strategies include **I**mplementation and enforcement of laws, **N**orms and values, **S**afe Environments, **P**arent and caregiver support, **I**ncome and economic strengthening, **R**esponse and support services, and **E**ducation and life skills (3). The concrete recommendations included in the INSPIRE handbook mark a defining step forward for ending violence against children. Further evidence around the cost-effectiveness of INSPIRE strategies could help stakeholders to prioritise resources to strategies that are likely to have the greatest return on investment (4).

Economic evaluations of violence prevention interventions remain relatively uncommon, particularly in low- and middle-income countries (5–8). Experimental studies are recommended for evaluating cost-effectiveness (9–11). However, the cost and complexity of such studies, especially when more than one intervention is under consideration, means that another methodology may be better suited to filling the urgent need for estimates of cost-effectiveness. Decision analytic modelling is a much cheaper alternative approach for generating estimates of cost-effectiveness that uses data from secondary sources including published trials and observational studies (7,9). By modelling different scenarios, this approach is also suited to generating early estimates of cost-effectiveness for novel interventions that have not yet been implemented. The validity and transferability of findings from decision analytic models is often limited by the quality of published evidence that is available in the literature.

Decision-analytic modelling provides an opportunity to use evidence from published studies to generate early estimates of cost-effectiveness for INSPIRE strategies (12,13). A previous study to guide the choice of ‘best-buy’ INSPIRE strategies in South Africa found that across seven INSPIRE-aligned protective factors food security, caregiver supervision, and positive caregiving are likely to be promising targets for addressing multiple adolescent violence outcomes simultaneously (14). Consistent with positive youth development theories, these three protective factors were also found to combine additively, such that experiencing two or three of them together was associated with a significantly lower probability of experiencing multiple forms of violence, compared to experiencing one of them alone (14). Findings from an experimental trial in Tanzania also support the additive combination of parenting support and economic strengthening, compared to either of the two interventions alone (15).

Identification of positive and supervisory caregiving and food security as promising intervention targets for preventing multiple violence outcomes marks a step forward towards helping stakeholders identify INSPIRE strategies that are likely to be particularly effective (16). However, to inform resource-prioritisation further work is needed to select candidate interventions that could be used to strengthen food security, and/or positive and supervisory caregiving, and draw together sufficient data to compare the expected effects of selected interventions against their associated cost. Aiming to do this, we had three objectives in this study 1) Select expert- and evidence-based interventions for reducing violence via improving food security, caregiver supervision, or positive caregiving in South Africa; 2) Draw together cost and effectiveness data relating to selected interventions from a variety of high-quality sources; 3) Combine this data in a decision-analytic model to estimate the cost-effectiveness of selected interventions.

## Methods

This economic modelling study builds on a non-randomised analysis of seven INSPIRE-aligned protective factors for violence victimisation in South Africa (14). The analysis found that food security, caregiver supervision, and positive caregiving were associated with lower odds of physical abuse, sexual abuse, emotional abuse, community violence victimisation, youth lawbreaking, and transactional sex. It also found that these protective factors combined additively, such that compared to experiencing a single protective factor alone, experiencing two, or all three simultaneously was associated with significantly lower odds of violence outcomes.

Our methodological approach of modelling the impact of interventions on violence outcomes had three steps. First, we consulted with regional experts in violence prevention to identify the best candidate interventions for reducing violence via improving food security, caregiver supervision, positive caregiving in South Africa. Second, we drew together cost and effectiveness data relating to the selected interventions from a variety of high-quality sources including published literature and online survey data. Third we combined these data in a decision-analytic model to estimate the total DALYs averted, cost, and cost-effectiveness of our selected interventions. Our choice to use DALYs averted was based on generating a single estimate of cost-effectiveness for selected interventions. We include full details of data-sources and assumptions throughout the analysis and report our study using the Consolidated Health Economic Evaluation Reporting Standard (CHEERS) checklist, S1 Appendix (17,18).

### Study setting

The study setting was Mpumalanga Province, where intervention costs and adolescents’ experience of violence are expected to lie between the other two provinces where data were collected for the non-randomised analysis of seven INSPIRE-aligned protective factors for reducing adolescent violence in South Africa; the poorer Eastern Cape Province, and richer Western Cape Province (14,19).

### Target population

The target population was households with a monthly household income per capita below the South African food poverty line of United States Dollar (USD) 39 and an adolescent aged 11-19 years (20–22).

### Hypothesised interventions for violence prevention

We consulted with experts in violence prevention to identify the best candidate interventions for reducing adolescent violence via improving food security, caregiver supervision, positive caregiving in South Africa. Our focus on these protective factors was based on evidence of their association with lower probability of three or more types of violence against adolescents in South Africa (14). Criteria for candidate interventions were that (i) there should be existing or early implementation within South Africa; and ii) evidence supporting their impact on either food security or positive and supervisory caregiving in Southern Africa. Key details on intervention implementation, staffing, and duration are provided in S2 Appendix (14).

#### Intervention 1: Outreach intervention to link households that are eligible but not receiving South Africa’s Child Support Grant (CSG) [Grant outreach]

It is estimated that around 20% of children eligible for anti-poverty social grants in South Africa do not access this support (23,24). Regional evidence supports the effectiveness of cash grants to address food security (25,26); and evidence from South Africa supports the potential of community initiatives to link eligible households to social grants (27). Therefore, in our analysis, we modelled the impact of a community outreach intervention implemented over 17 months during which paraprofessional social workers would actively liaise with community leaders and networks to screen for households that are eligible for the CSG in the community. Eligible households not already receiving the CSG would then be supported to access it.

#### Intervention 2: Parenting support intervention based on WHO/UNICEF’s Parenting for Lifelong Health (PLH) programme [Parenting Support]

Across South Africa, parenting support programmes are offered by a variety of non-profit organisations, including in Mpumalanga as part of the Mothers2Mothers (M2M) Children and Adolescents are My Priority (CHAMP) project (28,29). Evidence also supports the potential of parenting programmes for improving caregiver supervision in South Africa (30). In our analysis, we thus modelled the roll-out of a parenting support intervention akin to WHO’s Parenting for Lifelong Health Teen (PLH Teen), structured as a 14-session intervention for small family groups that are recruited by self or community-referral and using the two screening questions: ‘do you and your teen argue and shout a lot?’ and ‘do you sometimes end up hitting your teen when things are really stressful?’ (30,31).

#### Intervention 3: Integrated parenting support intervention plus component to link households that are eligible but not receiving South Africa’s CSG [Parenting support plus grant linkage]

In this combined scenario, we sought to explore the additive benefit of an intervention acting on two protective factors. We modelled a parenting-plus intervention structured similarly to the 14-session PLH Teen programme but including one additional session during which facilitators would assess families for their eligibility for inclusion in the CSG. Families found to be eligible but not receiving the CSG would be linked up with relevant social services to support them in accessing the grant. This parenting support plus grant linkage intervention would be distinct from the community grant outreach intervention as it would be focused on families that attend the parenting support intervention rather than the broader community.

### Choice of model

To estimate the cost-effectiveness of selected interventions, we constructed a probability tree model that modelled i) the effect of selected interventions on intermediary protective factors, and ii) the corresponding effect of the estimated improvement in intermediary protective factors on adolescent violence victimisation, Fig 1. Mixed evidence for whether group-based parenting support improves positive caregiving, meant that we did not consider parenting support or parenting support ‘plus’ grant linkage acting via this intermediary protective factor. Furthermore, our choice to measure intervention effectiveness in disability-adjusted life years (DALYs) averted meant that we could only consider violence outcomes with evidence for their attributable DALYs in our model. Of the six violence outcomes investigated in the non-randomised analysis of seven INSPIRE-aligned protective factors in South Africa, this included emotional abuse, physical abuse, and sexual abuse; but excluded youth lawbreaking, community violence victimization, and transactional sex (14). Finally, no evidence for an association between caregiver supervision and sexual abuse in the analysis of INSPIRE aligned protective factors, meant that we did not consider parenting support or parenting support ‘plus’ grant linkage acting on this outcome (14). With these considerations, we modelled community grant outreach acting on physical, emotional, and sexual abuse via food security, parenting support acting on physical and emotional abuse via caregiver supervision, and parenting support plus grant linkage intervention acting on the same outcomes via both food security and caregiver supervision.

**Fig 1.**
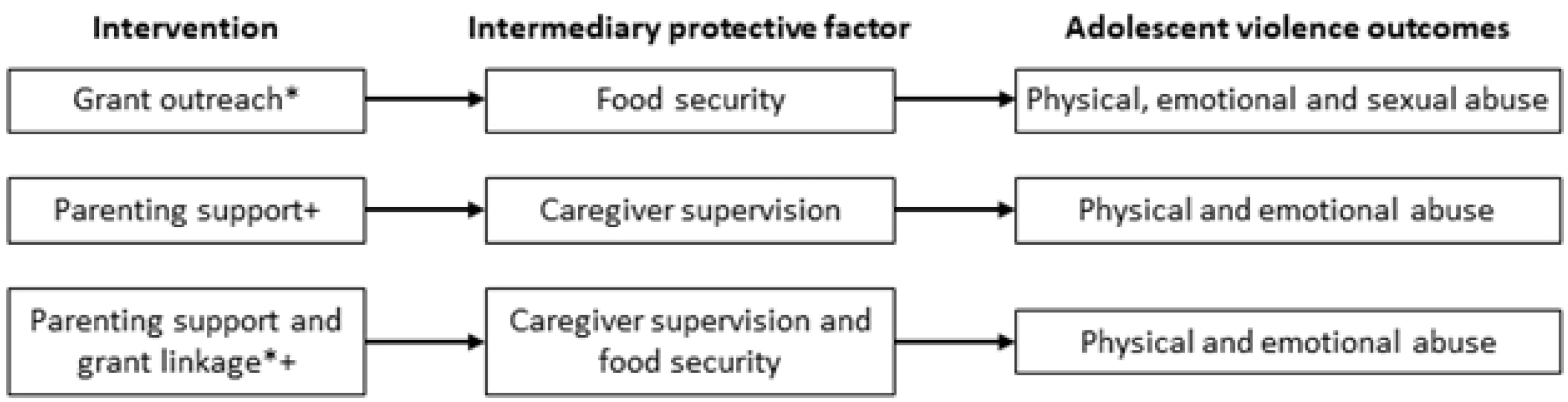
Graphical representation of our probability tree model for estimating the effectiveness of interventions on adolescent violence outcomes. *Estimate synthesized across two studies examining the impact of grants on household food security in sub-Saharan Africa and crosschecked using data from South Africa’s 2018 General Household Survey. +Estimate obtained from the primary analysis of a randomised evaluation of PLH Teen in South Africa.

### Data sources

Estimates of the percentage of adolescents in Mpumalanga living below the food poverty line were derived from South Africa’s 2018 General Household Survey (32). Estimates of the prevalence of adolescent violence outcomes were obtained from the non-randomised analysis of seven INSPIRE-aligned protective factors for adolescent violence victimisation in South Africa (14).

#### Intervention effects

Estimates for intervention effectiveness were drawn from the academic literature, S3 Appendix. For community grant outreach, we generated a pooled estimate for the effect of cash transfers on food security by meta-analysis of two studies identified using a rigorous literature review, S4 Appendix (25,26). For parenting support, we used effect estimates for caregiver supervision taken from the original randomised evaluation of the PLH Teen intervention in South Africa (30). For parenting support plus grant linkage, we used our pooled estimate for the effect of cash transfers on food security and our effect estimate for caregiver supervision from the PLH Teen evaluation. Finally, for the effect of enhanced levels of protective factors on adolescent violence victimisation we used estimates from the non-randomised analysis of seven INSPIRE-aligned protective factors for reducing adolescent violence in South Africa (14).

#### Intervention costs

All three interventions were costed from the provider’s perspective using the ingredients method. Unit costs and quantities for parenting support and parenting support plus grant linkage interventions were based on data from the PLH Teen trial in South Africa, and PLH implementation in Thailand and Tanzania. We were unable to find cost data on an intervention similar to the modelled grant outreach intervention, so unit costs were based on the PLH Teen trial and unit quantities on expert consultation.

### Data management

All costs captured before 2021 were adjusted for inflation and converted to USD using the conversion rate of USD 1 = South African Rand 14.93.

### Data analysis

#### Modelling the effectiveness of interventions

We used our probability tree model to calculate the number of cases of emotional, physical, and sexual abuse that each intervention would avert per year based on average, upper, and lower effect sizes. According to WHO-CHOICE guidelines, we modelled the number of cases of violence averted by selected interventions over 10 years at full implementation. Assumptions about the modelled annual decay in intervention effectiveness and exit rates are included in S5 Appendix. We converted estimated subtotals of cases averted into DALYS averted and summed across them to estimate total DALYS averted for each intervention. DALYs attributable to physical, emotional and sexual abuse were calculated from the only known evaluation of the economic consequences of violence against children in South Africa, and estimates of the absolute prevalence of each form of violence in South Africa, S6 Appendix (6,33,34).

#### Modelling the cost of interventions

Intervention costs were calculated over 10 years applying an annual discount rate of 4% (35). For all three interventions, we considered a ‘routine service delivery’ and ‘trial-based’ cost scenario. These two scenarios targeted the same number of families but varied in staff salaries, with the later providing significantly higher salaries, and budgeting for venue hire, printed training materials, and communication with participants via mobile phones. Routine service delivery costs were based on daily salary rates equivalent to South Africa’s average monthly earnings in March 2021, while trial-based costs were based on daily salary rates three times higher than this (36).

#### Modelling the cost-effectiveness of interventions

We calculated average cost-effectiveness ratios (ACERs) for community grant outreach and parenting support, by dividing intervention total costs by total DALYs averted. We calculated an incremental cost-effectiveness ratios (ICERs) for parenting support plus grant linkage, by dividing the cost of adding one additional session to link eligible households to the CSG by the additional DALYS averted by this approach relative to standalone parenting support (9,18). Estimated cost-effectiveness ratios and their upper and lower bounds were evaluated against a South African-specific willingness-to-pay (WTP) threshold estimated by Stacey and Edoka (37).

We calculated cost-effectiveness ratios for all three of our interventions under routine service delivery and trial-based costings, as well as at two estimates of the prevalence of adolescent violence outcomes. These were ‘population-average’ prevalence, which was equal to the rates observed in our source reference; and ‘high’ prevalence, which was equal to double the rates observed in our source reference (14). The rationale for modelling a high-prevalence of violence was that households targeted by our grant-outreach intervention (i.e., eligible but not receiving the CSG), and our parenting support and parenting support plus grant linkage interventions (i.e., screened for regular arguments and/or physical abuse against children) would experience significantly higher than average levels of vulnerability and adolescent violence victimisation.

#### Robustness checks

First, we evaluated the robustness of our findings to use of South Africa’s GDP per capita for 2021 as our willingness to pay threshold (18,38). Second, given the absence of a formal evaluation of the effect of South Africa’s CSG on a subjective measure of food security in the literature, we also checked the robustness of findings for the cost-effectiveness of community grant outreach, and parenting support plus grant linkage to a country-specific estimate for the relationship between income and food security estimated using South Africa’s 2018 General Household Survey (39). Further details of this secondary analysis are included in S7 Appendix.

### Ethics

The study used secondary data published in the public domain throughout the analysis.

## Results

Details on the demographics of Mpumalanga Province, and the target population for interventions are summarized in Table 1. We observed that one out of three households with an adolescent in Mpumalanga live below the food poverty line, corresponding to 212,000 adolescents. We estimated that approximately 66,000 adolescents living in 64,300 households would benefit from the grant outreach intervention. We also estimated that over 10 years of intervention implementation, roughly 70,000 adolescents living in 68,000 households would benefit from the parenting support or parenting support plus grant linkage interventions.

**Table 1.** Core model variables.

| Variable | Value | Assumption | Source |
| --- | --- | --- | --- |
| **Demographics of Mpumalanga province** |  |  |  |
| Total population | 4,523,000 | - | Mid-year population estimate, Statistics South Africa |
| Total households | 1,269,000 | Average household size = 3.5 | South Africa Community Survey, 2016 |
| Total adolescents | 591,000 | % of adolescents = 19.3 | Demographic Profile of Adolescents in South Africa, Statistics South Africa (40) |
| Total adolescents living below the food poverty line | 212,000 | % of adolescents living below the poverty line = 34.9 | South Africa GHS 2018 (32) |
| **Target population for a grant outreach intervention** |  |  |  |
| Eligible adolescents not receiving the CSG | 94,500 | % of eligible households with children not receiving the CSG = Average number of adolescents (11-18 years) per household with children | UNICEF, 2016 (24)<br>South Africa GHS, 2018 (32) |
| Ideal coverage for the target population | 100% | The intervention aims to link up all eligible households and adolescents |  |
| Expected number of adolescent beneficiaries | 66,000 | 70% success rate in linking eligible children to CSG | Expert consultation; Thurman et al (27) |
| Expected number of household beneficiaries | 64,300 | Number of adolescents per household = 1.03 | Calculated |
| **Target population for parenting support/ parenting plus grant linkage interventions** |  |  |  |
| Ideal coverage for the target population per annum | 5% | Intervention targeted towards low-income households and considered resource requirements and scale-up observed in other countries | Consultation with experts and implementation managers |
| Expected number of adolescent beneficiaries | 70,000 | 90% expected uptake rate representing intervention effectiveness in enrolling targeted households | Over 95% compliance was recorded during the PLH Teen trial (30). Lowered to 90% that aims to account for expected lower compliance in a real-world setting |
| Expected number of household beneficiaries | 68,000 | Number of adolescents per targeted household that takes up intervention = 1.03 | Calculated |

Abbreviations: CSG, child support grant; GHS, General Household Survey.

### Estimated intervention effects

Estimates for the expected effect of interventions via respective pathways are reported in Table 2.

**Table 2.** Estimated effectiveness of modelled interventions.

|  | Average effect size (ppts) | Lower; Upper effect size (ppts) | Source |
| --- | --- | --- | --- |
| Intervention 1. Community grant outreach |  |  |  |
| Effect on food security | +14.41 | +4.29; +23.80 | Review of studies examining the impact of social grants on food security in Sub-Saharan Africa (26,41), Cross-validated by a cross-sectional multivariable analysis examining the association between monthly household income per capita and food security using South Africa GHS, 2018 (32) . |
| Effect of enhanced food security on adolescent violence |  |  |  |
| Sexual abuse | -0.54 | -0.86; -0.16 | Longitudinal multivariable analysis examining the relationship between food security and multiple violence in South Africa (14) |
| Emotional abuse | -0.86 | -1.40; -0.25 |  |
| Physical abuse | -1.66 | -1.66; -0.30 |  |
| Intervention 2: Parenting support |  |  |  |
| Effect on parental monitoring | +1.21 | +0.32; +2.09 | Cluster randomised controlled trial evaluating PLH Teen parenting support intervention(30) |
| Effect of enhanced parental monitoring on adolescent violence |  |  |  |
| Sexual abuse* | - | - | Longitudinal multivariable analysis examining the relationship between caregiver supervision and multiple violence outcomes in South Africa (14) . |
| Emotional abuse | -0.71 | -0.73; -0.69 |  |
| Physical abuse | -0.97 | -0.99; -0.95 |  |
| Intervention 3: Parenting support plus grant linkage |  |  |  |
| Effect on food security | +14.41 | +4.29; +23.80 | Equivalent to the exact estimate of the impact of Interventions 1 and 2 on protective factors. |
| Effect on parental monitoring | +1.21 | +0.32; +2.09 |  |
| Effect of enhanced food security and parental monitoring on adolescent violence |  |  |  |
| Sexual abuse* |  |  | Longitudinal multivariable analysis examining the relationship between caregiver supervision, and multiple violence in South Africa (14). Estimated effects are greater than for parenting support intervention alone as they consider the additive effects of experiencing enhanced food security and parental monitoring together. |
| Emotional abuse | -0.95 | -0.85; -0.97 |  |
| Physical abuse | -0.94 | -0.90; -0.98 |  |

\*Violence outcome was not considered in the cost-effectiveness model, as there was no evidence that it was significantly associated with caregiver supervision in (9). Abbreviations: ppts, percentage points; GHS, General Household Survey.

#### Grant outreach

We estimated that linking eligible households to the CSG would increase the household’s probability of food security by +14.41ppts (+4.29; +23.80). Based on our model, a grant outreach intervention would be expected to reduce the probability of sexual abuse by - 0.54ppts (−0.86; −0.16), emotional abuse by −0.86ppts (−1.40; −0.25), and physical abuse by - 1.66ppts (−1.66; −0.30).

#### Parenting support

The results of the PLH Teen intervention suggest that the parenting support intervention would increase caregiver supervision by 1.21ppts (0.32; 2.09ppts). Based on our model, a parenting support intervention would be expected to reduce the probability of emotional abuse by −0.71ppts (−0.73; −0.69ppts), and physical abuse by −0.97ppts (−0.99; - 0.95ppts) through improvements in caregiver supervision alone.

#### Parenting support plus grant linkage

When considering the combination of parenting support and linkage to the CSG for increasing households’ probability of food security and caregiver supervision, we estimated that parenting support plus grant linkage would reduce the probability of emotional abuse by −0.95ppts (−0.85; −0.97) and physical abuse by −0.94ppts (−0.90; −0.98).

### Estimated intervention costs

Cost estimates for interventions are reported in Table 3.

**Table 3.** Summary of the estimated cases of violence averted and costs per adolescent of intervention scenarios over ten years implementation period.

| Intervention | Total adolescent beneficiaries | Averted cases of violence (Low; Upper bound) |  | Total cost per adolescent, USD |  | Total costs provincial, USD |  |
| --- | --- | --- | --- | --- | --- | --- | --- |
|  |  | Population-average prevalence of violence | High prevalence of violence* | Routine service delivery scenario | Trial-based scenario | Routine service delivery scenario | Trial-based scenario |
| Grant outreach | 66,000 | 5,800<br>(1,800; 9,500) | 11,600<br>(3,500; 19,000) | 61 | 111 | 4,035,000 | 7,300,000 |
| Parenting support | 70,000 | 3,700<br>(940; 6,000) | 7,700<br>(1,900; 12,000) | 36 | 215 | 2,530,000 | 15, 075,000 |
| Parenting support plus grant linkage | 70,000 | 4,800<br>(1,700; 10,200) | 9,600<br>(3,500; 20,400) | 37 | 223 | 2,616,700 | 15,603,000 |

Abbreviations: USD, United States Dollar.

#### Grant outreach

With routine service delivery and trial-based budget scenarios, community grant outreach was estimated to cost USD 61 and USD 111 per adolescent; and USD 4,035,000 and USD 7,300,000 at the provincial level, respectively.

#### Parenting support

With routine service delivery and trial-based budget scenarios, parenting support was estimated to cost USD 37 and USD 215 per adolescent beneficiary, and USD 2,530,000 and USD 15,075,000 over 10 years at the provincial level (USD 275,700 and 1,640,000 annually), respectively. The major cost drivers were staff salaries (54% and 80% of total costs for trial-based and routine service delivery, respectively), and food for participants and facilitators (21% and 0% of total costs under the trial-based and routine service delivery, respectively).

#### Integrated parenting support plus grant linkage

With routine service delivery and trial-based budget scenarios, integrated parenting support plus grant linkage was estimated to cost USD 37 and USD 223 per adolescent, and USD 2,617,000 and USD 15,603,000 over 10 years at the provincial level, respectively.

### Intervention cost-effectiveness

Intervention ACERs are summarised against a South African specific WTP threshold for each of the combinations of population-average versus high prevalence of violence and routine service delivery and trial-based intervention costs in Fig 2. An intervention is considered cost-effective if its ACER lies below the WTP threshold. The further an ACER lies to the right of the plane while remaining below the WTP threshold the higher its indicated cost-effectiveness.

**Fig 2.**
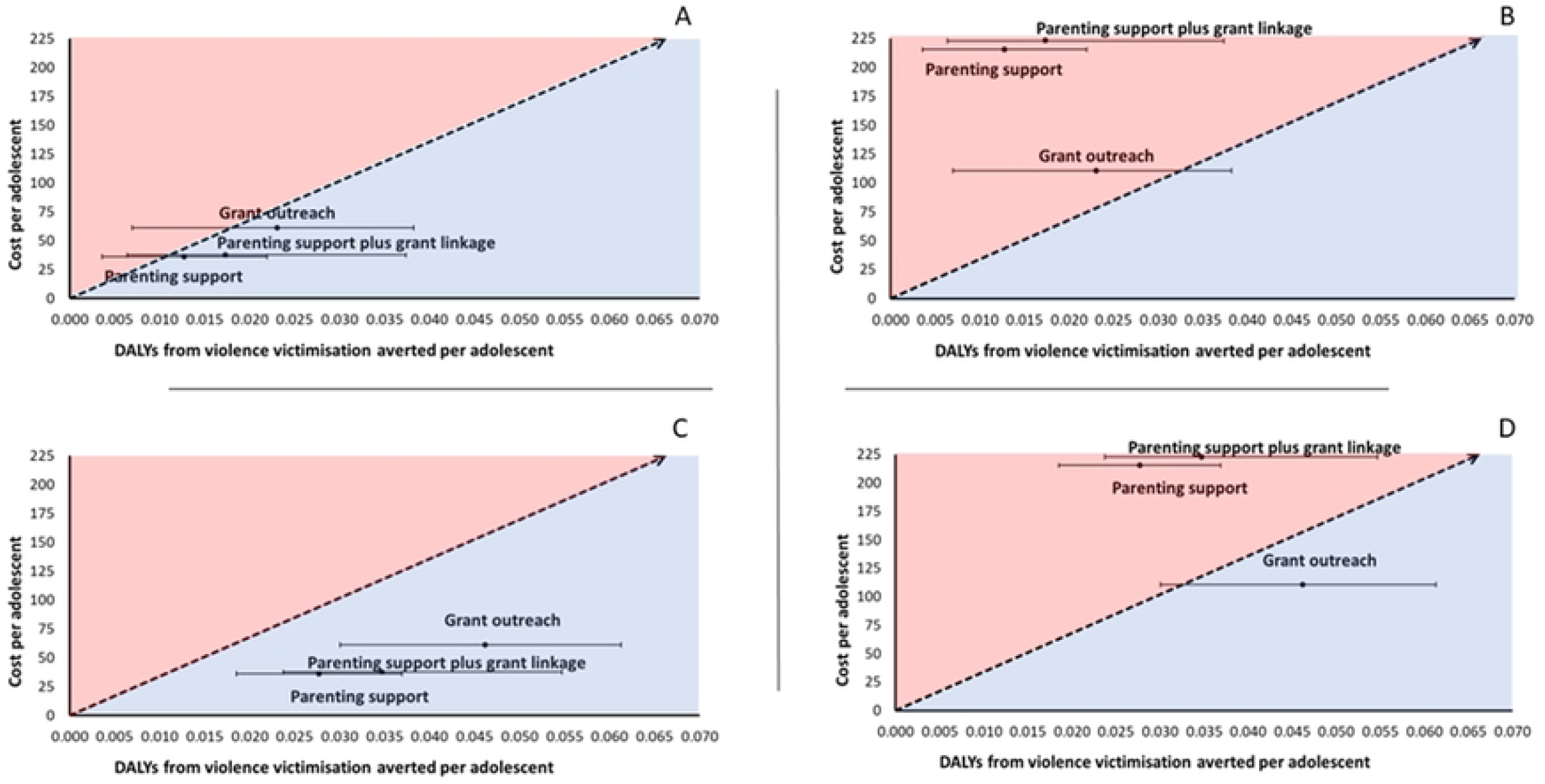
Cost-effectiveness plane scatter plots of costs per adolescent over effectiveness (in DALYs averted) for four scenarios at A) Population-average prevalence of violence and routine service delivery costs, B) Population-average prevalence of violence and trial-based costs C) high prevalence of violence and routine service delivery costs, and D) high prevalence of violence and trial-based costs. Diagonal lines indicate an evidence-based SA-specific willingness to pay threshold for health interventions estimated by Edoka et al. Strategies become increasing cost-effective as they lie to the north easternmost part of the cost-effectiveness plane below the willingness to pay threshold. Strategies above the line are not cost-effective at the stated threshold. The values underlying this graph are reported in S8 Appendix. Abbreviations: DALYs, Disability-Adjusted Life Years; USD, United States Dollar.

#### For a population-average prevalence of violence

Under a routine service delivery cost scenario, ACERs for community grant outreach and parenting support were both below the WTP threshold at USD 2,650 and USD 2,830 per DALY averted respectively. Integrated parenting support plus grant linkage had an ICER of USD 271 per DALY averted compared to the parenting support intervention.

Under a trial-based cost scenario, ACERs for community grant outreach and parenting support were both above the WTP threshold at USD 4,800 and USD 16,880 per DALY averted respectively. The integrated parenting support plus grant linkage was also not cost-effective with an ICER of USD 12, 850 per DALY averted.

#### For a high prevalence of violence

Under a routine service delivery cost scenario, ACERs for community grant outreach and parenting support were both below the WTP threshold at USD 1,330 and USD 1,310 per DALY averted. The integrated parenting support plus grant linkage intervention had an ICER of USD 177 per DALY averted compared to the parenting support intervention.

Under a trial-based cost scenario, the ACER for community grant outreach was below the WTP threshold at USD 2,400 per DALY averted, but the ACER for parenting support was above the WTP threshold at USD 7,780 per DALY averted. The ICER for integrated parenting support plus grant linkage was USD 6,430 per DALY averted and above the WTP threshold.

### Sensitivity analysis and robustness check

When using the WHO CHOICE threshold based on South African GDP per capita instead of a South Africa-specific WTP threshold, results were the same except that at a population-average prevalence of violence, under trial-based costing, the ACER for community grant outreach was below the WTP threshold, S8 Appendix. Substituting our pooled estimate for the effect of cash transfers on food security with an estimate of the relationship between income and food security from the South Africa’s 2018 General Household Survey, the findings for parenting support plus grant linkage remain consistent, but community grant outreach ceases to be cost-effective at for the population-average prevalence of violence and routine service delivery cost scenario, or a high prevalence of violence and trial-based cost scenario, S9 Appendix.

## Discussion

We find that decision-analytic modelling is a valuable and low-cost approach for assessing the cost-effectiveness of three expert- and evidence-informed interventions aimed at reducing adolescent violence in South Africa. Our model indicates that implementing each of our three selected interventions using a routine service delivery model is likely to be cost-effective for reducing emotional, physical, and sexual abuse against adolescents. This finding is robust to all of the modelled combinations of low, average, and high intervention effectiveness with an average or high prevalence of violence, except for the combination of low intervention effectiveness and an average prevalence of violence in the target population. Across the three interventions, the most cost-effective option is likely to be the community grant outreach intervention. Thereafter, the integrated parenting support plus grant linkage intervention is estimated to be more cost-effective than the parenting support intervention alone. We also find that if our three selected interventions are implemented using trial-based costings, parenting support and parenting plus grant linkage support are unlikely to be cost effective, and community grant outreach is only likely to be cost-effective assuming average to high intervention effectiveness, and higher than average prevalence of violence in the target population.

Our study provides early evidence around the cost-effectiveness of a community outreach intervention to support household’s access to social grants if they are eligible, but not yet receiving them. The cost of the proposed grant outreach intervention per adolescent is equivalent to the value of three household child support grants and falls on the lower end of the scale of home-visiting interventions in high-income settings (42). While our results are promising, evidence around the effectiveness of cash transfers alone for preventing adolescent violence victimisation remains limited, and this is an important area for further research (43,44).

There are still limited economic evaluations of adolescent violence prevention interventions in low- and middle-income countries, particularly of large-scale interventions (45,7). Our findings around the cost-effectiveness of parenting support implemented using routine-delivery costs matches a previous economic evaluation of the original PLH Teen intervention in South Africa that also modelled a similar ‘lean’ scenario (6). An assumption in both of these studies is that the modelled routine-service delivery intervention is just as effective as the intervention implemented using trial-based costings. However, is increasingly supported by evidence that when implemented in the real-world at much lower cost PLH Teens is just as effective as that observed in experimental studies (46). The increasing roll-out and scale-up of parenting support programmes across sub-Saharan Africa will present further opportunity to validate this in the future (47).

Unlike for routine service delivery, our findings around the cost-effectiveness of trial-based delivery of parenting support do match the previous economic evaluation of PLH. Possible explanations for this discrepancy may be our focus on the indirect effects of parenting support via caregiver supervision alone rather than the total effect via all possible pathways. Other impact pathways have been found to include improved caregiver mental health, caregiver alcohol/drug avoidance, and improved economic welfare (48). Another explanation could be the previous evaluation modelled a higher prevalence of adolescent violence victimisation among families targeted by the PLH Teen intervention than we considered in either our population-average or high prevalence of violence scenarios(49).

While there is growing evidence on the enhanced effectiveness of combining multiple interventions with a ‘plus’ approach, there is still very little evidence around the cost-effectiveness of these interventions (50,51). Our findings of the incremental cost-effectiveness of combining an additional grant linkage session with PLH Teen supports the added value of integrated services for reducing adolescent violence victimisation. Building on this, future research should consider other possible approaches for combining parenting support with an economic strengthening intervention to simultaneously act via caregiver supervision and food security (52–54).

Our modelling study enabled us to consider important questions around the design of three interventions for reducing violence against children. These included how best to build on existing social policies such as South Africa’s CSG, the resources needed for implementing interventions at scale, and approaches for combining two interventions to simultaneously promote food security and caregiver supervision (3). Strengths of our study include its significantly lower cost compared to running a randomised evaluation of the three evaluated interventions, and also our use of a widely applicable methodology that could be used to generate similar evidence for other interventions and/or settings. However, our study also had limitations. While many of our model parameters were drawn from high-quality randomised studies, estimates of association between protective factors and violence outcomes were based on observational research and may be affected by sources of bias associated with this type of research design including unmeasured confounding (14). Gaps in the academic literature also meant that we had to make assumptions about some of the parameters in our model, for example the post-intervention decay in effectiveness of cash grants and parenting programmes. Several methodological choices also mean that our estimates of cost-effectiveness are likely to be conservative. These include our focus on DALYs averted for measuring cost-effectiveness, which while allowing us to generate a single estimate of cost-effectiveness for selected interventions, also meant we could not consider outcomes that lacked estimates of avoidable DALYs in our model including community violence victimisation, youth lawbreaking, or transactional sexual exploitation. Also, our focus on the impact of interventions via food security and caregiver supervision alone did not consider the full complement of protective factors via which selected interventions could act on adolescent violence outcomes. There is growing evidence that parenting support is also likely to act on adolescent violence victimisation via improving caregiver mental health, caregiver alcohol/drug avoidance, and improved economic welfare (48,55,56). Due to a lack of evidence in the wider literature, we were also unable to consider the wider societal benefits of preventing adolescent violence victimisation. Such benefits might include reduced health service use, social service use, and court case time. Finally, all three of the interventions considered in this study are likely to have benefits for adolescent wellbeing beyond reducing violence victimisation, for example by boosting school enrolment, or promoting mental health. Impacts across these additional domains of wellbeing should be considered for accurately valuing intervention cost-effectiveness in the future. Doing so may also support cross-sectoral buy-in from multiple government departments including health, education, and social development (57).

In South Africa, 40% of young people are estimated to experience some form of sexual, physical, or emotional abuse in their lives (58). Reviewing and analysing the literature on violence against children in South Africa, our study provides novel and policy-relevant evidence for much needed action to reduce child abuse. Our findings suggest that all three of our selected interventions are likely to be cost-effective so long as they are implemented using routine service delivery rather than a trial-based costing. The study also provides a comprehensive summary of evidence gaps that should be investigated as a priority for further informing the scale-up of interventions in this field. These include limited estimates of the prevalence of adolescent violence victimisation among the most vulnerable groups, the effects of cash transfers on adolescent violence victimisation or the mechanisms via which they act on this outcome, and the wider benefits of reducing violence against children to communities and society. Future attempts to quantify the cost-effectiveness of interventions like community grant outreach and parenting support studies should also account for their spill over effects on other areas of adolescent wellbeing including education and health.

## Conclusion

In conclusion, we find that investments in community grant outreach, and parenting support interventions are likely to be cost-effective for preventing violence against adolescents when these programmes are implemented as routine services. Adding a grant linkage component to parenting support is likely to enhance the cost-effectiveness of this intervention and supports the value of integrated services to accelerate progress in preventing violence against children. Further research would strengthen the accuracy of our model estimates.

## Data Availability

All relevant data are within the paper and its supporting information files.

## Acknowledgement

We thank Dr Jamie Lachman for providing us with costing data from past implementation of parenting programmes. We thank Dr Yulia Shenderovich for providing data from the parenting for Lifelog Health (PLH Teen) trial. We also thank Gloria Khoza and Mpume Danisa from UNICEF South Africa and Clown without Borders South Africa respectively, whose experience in scaling up parenting programmes in South Africa provided valuable insight into the intervention scenarios during the early phases of this work.

## Supporting Information

**S1 Appendix**. CHEERS Checklist.

**S2 Appendix**. Description of hypothesised interventions, their duration, and staffing structure.

**S3 Appendix**. Summary of data sources used to estimate the effectiveness and cost of selected interventions.

**S4 Appendix**. Synthesis of the effect of Cash Grants on Household Food Insecurity Access Scale (HFIAS) from selected studies in Sub-Saharan Africa.

**S5 Appendix**. Assumptions about the modelled annual decay in intervention effectiveness and exit rates.

**S6 Appendix**. Estimated DALY per case of physical, emotional, and sexual abuse.

**S7 Appendix**. Sensitivity analysis of the effect of cash transfers on food security in South Africa using data from the 2018 General Household Survey.

**S8 Appendix**. Average and Incremental Cost Effectiveness Ratios.

**S9 Appendix**. Cost-effectiveness plane scatter plots of costs per adolescent over effectiveness (in DALYs averted) for four scenarios with the SA GDP per capita as the willingness-to-pay threshold at: A) Population-average prevalence of violence and routine service delivery costs, B) Population-average prevalence of violence and trial-based costs, C) High prevalence of violence and routine service delivery costs, and D) High prevalence of violence and trial-based costs.

**S10 Appendix**. Cost-effectiveness plane scatter plots of costs per adolescent over effectiveness (in DALYs averted) for four scenarios with the SA-specific threshold per capita as the willingness-to-pay threshold at: A) Population-average prevalence of violence and routine service delivery costs, B) Population-average prevalence of violence and trial-based costs, C) High prevalence of violence and routine service delivery costs, and D) High prevalence of violence and trial-based costs.

